# Leveraging pathogen sequence and contact tracing data to enhance vaccine trials in emerging epidemics

**DOI:** 10.1101/2020.09.14.20193789

**Authors:** Rebecca Kahn, Rui Wang, Sarah V. Leavitt, William P. Hanage, Marc Lipsitch

## Abstract

**Introduction:** Advance planning of the design and analysis of vaccine trials conducted during infectious disease outbreaks increases our ability to rapidly define the efficacy and potential impact of a vaccine and inform public health response. Vaccine efficacy against infectiousness (VE_I_) is an important measure for understanding the full impact of a vaccine, yet it is currently not identifiable in many vaccine trial designs because it requires knowledge of the vaccination status of infectors. Recent advances in pathogen genomics have improved our ability to accurately reconstruct transmission networks. We aim to assess if augmenting classical randomized controlled trial designs with pathogen sequence and contact tracing data can permit these trials to estimate VE_I_.

**Methods:** We develop a transmission model with a vaccine trial in an outbreak setting, incorporate pathogen sequence evolution data and sampling as well as contact tracing data, and assign probabilities to likely infectors. We then propose and evaluate the performance of an estimator of VE_I_.

**Results:** We find that under perfect knowledge of infector-infectee pairs, we are able to accurately estimate VE_I_. Use of sequence data results in imperfect reconstruction of the transmission networks, biasing estimates of VE_I_ towards the null, with approaches using deep sequence data performing better than approaches using consensus sequence data. Inclusion of contact tracing data reduces the bias.

**Conclusion:** Pathogen genomics enhance identifiability of VE_I_ from individually randomized controlled trials, but imperfect transmission network reconstruction biases the estimates towards the null and limits our ability to detect VE_I_. Given the consistent direction of the bias, estimates obtained from trials using these methods will provide lower bounds on the true VE_I_. A combination of sequence and epidemiologic data results in the most accurate estimates, underscoring the importance of contact tracing in reconstructing transmission networks.

## Introduction

Vaccine trials conducted during epidemics of emerging infectious diseases provide an important opportunity to test the safety and efficacy of vaccine candidates. Increasing our ability to quickly and accurately understand the impact of a vaccine candidate in the urgent setting of an outbreak is critical for enhancing public health response. The use of the ring vaccination strategy in the *Ebola ça Suffit* trial during the 2013-2016 West African Ebola outbreak highlighted the importance of developing innovative designs for trials conducted during an ongoing outbreak.
^1^
 It also underscored the need to think through trial design and analysis strategies in advance in order to expedite the rollout of a vaccine trial once an outbreak starts and to identify the best methods for obtaining high quality efficacy estimates in outbreak settings.
^2^

Multiple components of vaccine efficacy can be estimated from a vaccine trial.
^3^
 Individually randomized controlled trials (iRCTs) estimate vaccine efficacy against susceptibility to infection (VE_S_), the direct effect of the vaccine on vaccinated individuals.
^3^
 If reducing susceptibility to infection is the only effect of the vaccine, then this measure, combined with information on contact network structure and pathogen transmission dynamics, can be used to estimate the total effect of a vaccination program, a combination of the direct and indirect (i.e. herd immunity) effects. Vaccine efficacy against infectiousness (VE_I_), the reduction in onward transmission from a vaccinated person who is infected compared to an unvaccinated infected person, is another important measure for understanding the impact of a vaccine.
^3^
 Even if a vaccine does not protect everyone who is vaccinated from getting infected, its impact on infectiousness for those who are vaccinated but nevertheless become infected plays a critical role in both outbreak dynamics and also cost-effectiveness of a vaccine program. The significance of understanding interventions’ effects on future transmission is exemplified by the efforts of HIV treatment-as-prevention programs to reduce patients’ viral loads to undetectable levels in order to prevent onward transmission.^
4,5
^

In order to estimate VE_I_, the vaccination status of infectors must be known. VE_I_ is therefore potentially measurable in household studies^
6,7
^ and partner transmission studies, such as HIV vaccine trials
^8^
 because in these settings, infector-infectee pairs can be identified (by assuming that household members or partners are the infectors), and thus the vaccination status of infectors is known. However, VE_I_ is not currently identifiable in population-level vaccine trials, such as those often conducted during an infectious disease outbreak, because the transmission network, and consequently the vaccination status of infectors, are typically unknown.

Recent advances in pathogen genomics have improved our ability to accurately reconstruct transmission networks.^
9–15
^ The West African Ebola epidemic and the ongoing COVID-19 pandemic have demonstrated our growing capacity to use sequence data in outbreak settings,^
16–21
^ and recent work has highlighted the potential for deep sequence data to add resolution to transmission networks.^
22–24
^ We aim to assess if augmenting classical randomized controlled trial designs with pathogen sequence data, as well as contact tracing data, would permit these trials to estimate VE_I_ by reconstructing transmission networks and identifying the trial status of infectors.

## Methods

We define θ as the risk ratio for becoming infected if one receives vaccine vs. control, or 1 – VE_S_, and Φ as the relative infectiousness of a vaccinated person who is infected compared to a control who is infected, or 1 – VE_I_. At the conclusion of a vaccine trial, the ratio of the proportion of people infected by vaccinated individuals to the proportion of people infected by controls is a product of both the vaccine’s effect on susceptibility to infection and its effect on infectiousness among those who are infected. With knowledge of who infected whom, using the ratio of infector vaccination status, we can therefore calculate VE_I_:

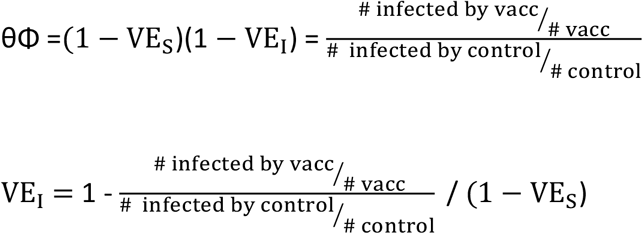

We simulate a compartmental network model of an outbreak, together with a vaccine trial, the details of which have been previously described.
^25^
 Individuals are grouped into communities, with many connections between individuals in the same community and fewer between individuals in different communities. Introduction of infection into the network occurs at a time-varying rate, and the disease natural history in the communities follows a stochastic susceptible, exposed, infectious, recovered (SEIR) model, with Ebola-like parameters (Table 1). Each individual has a daily probability of infection from their infectious contacts in the network. Individuals are enrolled into an iRCT, with 50% randomized to vaccine and 50% to control. The vaccine’s efficacy against susceptibility to infection is “leaky”, with 60% efficacy (VE_S_ = 0.60), meaning upon each exposure, the vaccine reduces a vaccinated individual’s chance of infection by 60%. The vaccine’s efficacy against infectiousness is 30%, meaning infectiousness among infected vaccinated individuals is 30% lower than among infected unvaccinated individuals (VE_I_ = 0.30). Table 2 shows the number of infections expected for each type of infector-infectee pair from the trial simulations.

**Table 1.**
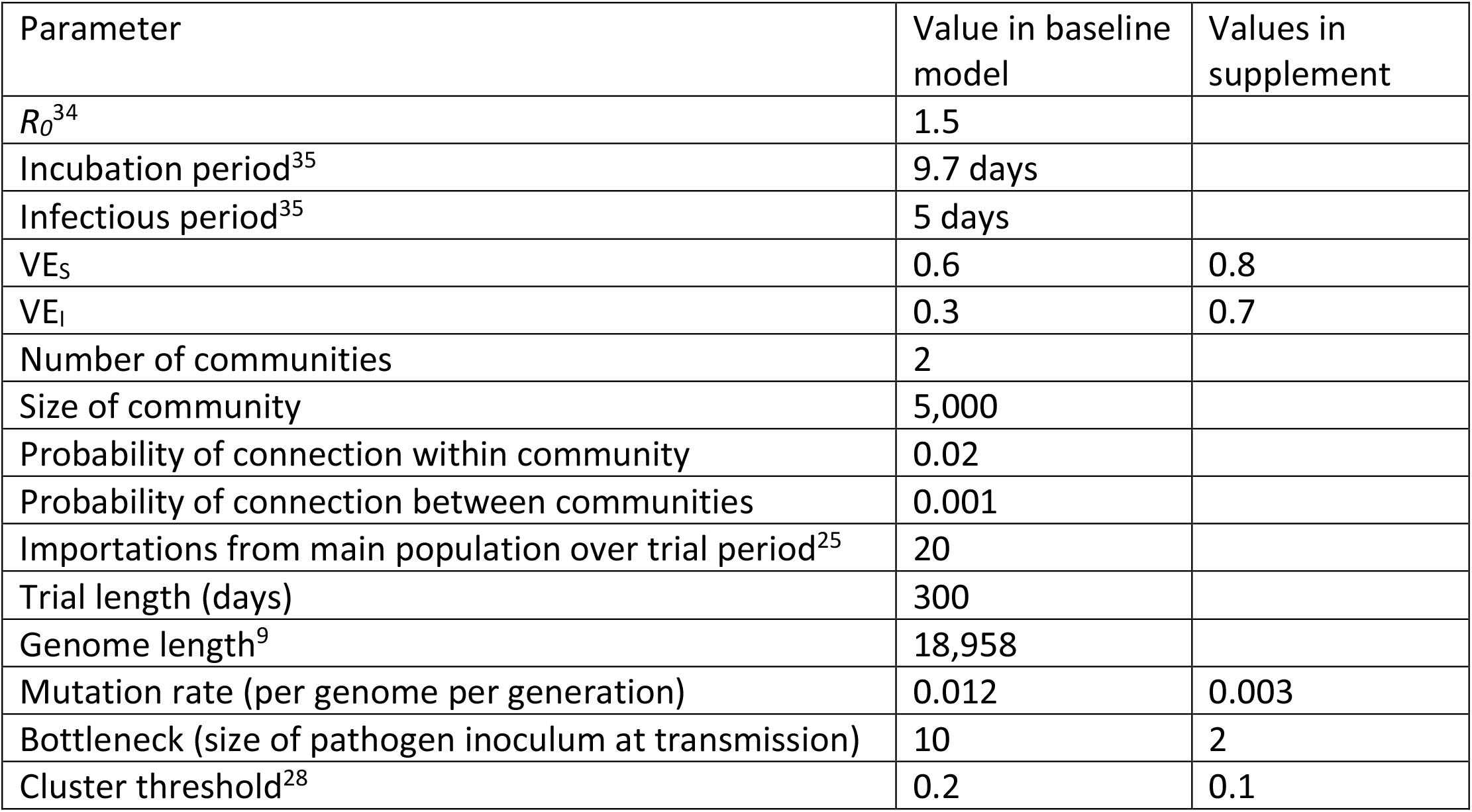

| Parameter | Value in baseline model | Values in supplement |
| --- | --- | --- |
| $R_0^{34}$ | 1.5 | |
| Incubation period<sup>35</sup> | 9.7 days |  |
| Infectious period<sup>35</sup> | 5 days |  |
| $VE_s$ | 0.6 | 0.8 |
| $VE_i$ | 0.3 | 0.7 |
| Number of communities | 2 |  |
| Size of community | 5,000 |  |
| Probability of connection within community | 0.02 |  |
| Probability of connection between communities | 0.001 |  |
| Importations from main population over trial period<sup>25</sup> | 20 |  |
| Trial length (days) | 300 |  |
| Genome length<sup>9</sup> | 18,958 |  |
| Mutation rate (per genome per generation) | 0.012 | 0.003 |
| Bottleneck (size of pathogen inoculum at transmission) | 10 | 2 |
| Cluster threshold<sup>28</sup> | 0.2 | 0.1 |

**Table 2.** A proportion *p* is randomized to vaccine and to control. In the absence of vaccination, a proportion *a* would become infected, and a proportion *2q* of all exposures to infection of participants would come from other trial participants (with *1–2q* external exposures). θ = 1 – VE_S_, or the risk ratio for becoming infected if one receives vaccine vs. control, and Φ = 1 – VE_I_, or the relative infectiousness of a vaccinated person who becomes infected to a control who becomes infected.

| *Infector (column)*<br>*Infectee (row)* | Vaccinee | Control | Any participant | Ratio of infectees |
| --- | --- | --- | --- | --- |
| Vaccinee | $apq\theta^2\Phi$ | $apq\theta$ | $apq\theta(1+\theta\Phi)$ | $\theta$ |
| Control | $apq\theta\Phi$ | $apq$ | $apq(1+\Phi)$ | |
| Any participant | $apq\theta\Phi(1+\theta)$ | $apq(1+\theta)$ | $apq(1+\theta)(1+\theta\Phi)$ | |
| Ratio of infectors | $\theta\Phi$ | | | |

To estimate VE_I_, we first make the unrealistic assumption of complete knowledge of the transmission network, with perfect ascertainment of who infected whom and their infection and recovery times. We then relax the assumption of perfect knowledge of who infected whom. Using the R package *seedy*,
^26^
 we incorporate pathogen evolution and sampling of both consensus and deep sequence data into the simulations, specifying parameters such as genome length, mutation rate, and bottleneck size (Table 1). As the choice of parameters, particularly mutation rate and bottleneck size, greatly impacts our ability to reconstruct transmission networks,
^22^
 we vary parameters across simulations to assess their impact on our ability to estimate VE_I_.

For each infectee we then assign a probability to each potential source of infection, based on comparisons of the sequence data for the candidate source(s) and each index case using four different approaches. In the first two approaches we use consensus sequence data. First, we assign probabilities to potential infectors based on the inverse of the genetic distance between the infectee and potential infectors. Second, we use a geometric-Poisson approximation of SNP distance to assign probabilities to potential infectors; this approach assumes genetically similar sequences are more likely to be infector-infectee pairs, while also accounting for mutation rate and times of infection.
^26^
 Third, we weight potential infectors by the number of rare variants (i.e. minority variants not seen in the consensus sequence that are rare in the population) they share with each infectee, which may be identified through deep sequence data and has previously been shown to provide additional resolution.
^22^
 Fourth, we combine the second and third approaches, using the consensus sequence data in the event that no shared minority variants for an infectee are identified through deep sequence data.
^22^
 For all four approaches, we then weight the probabilities identified through the sequence data by the probability of infection given the time of symptom onset of the infectee and potential infector(s) based on the serial interval distribution (i.e. the time between when an infector becomes symptomatic and their infectee becomes symptomatic).

Using each of these approaches, we then estimate the ratio of the number of cases infected by a vaccinated person to the number of cases infected by a control. We do this in three ways for each approach (see supplemental text 1 for more details). First, we weight each identified potential infector by the probability assigned to them and sum the probabilities by vaccination status. Second, we split the probabilities for each infectee into clusters based on the largest gap in probabilities between potential infectors.
^28^
 If the gap is larger than the specified threshold, we use the normalized probabilities from the infector(s) in the top cluster; otherwise we exclude that infectee from the analysis. Third, we use only the vaccination status of the most likely infector(s) for each infectee. Using the estimated ratio of the trial status of the potential infectors and the estimate of VE_S_ from the trial, we then estimate VE_I_, using the equation above. To incorporate the data from the network obtained through contact tracing efforts during an epidemic, we also conduct all of the approaches described above in a data set restricted to only potential infectors who are contacts of the infectees (i.e. connections in the network model).

We propose the following procedure for estimating the standard errors of the estimates under the approaches that perform best. For a given simulation and estimate of VE_I_, we obtain a bootstrap estimate of the standard error as follows. We first sample with replacement from the infected individuals. We then construct a bootstrapped data set using each infected individual from the sample and all of their potential infectors identified by the approach. We estimate VE_I_ from the bootstrapped data set and then repeat these steps 100 times. The standard deviation of the 100 bootstrapped estimates is the standard error of the VE_I_ estimate. This approach could be used with real data observed in a real trial and resembles the bootstrapping clusters approach (i.e., clusters are treated as units for resampling) for clustered data.
^29^
 Code is available: https://github.com/rek160/ADAGIO_WPA.

## Results

As expected, under perfect knowledge of the transmission network, VE_I_ is estimated correctly (median of 500 simulations: estimate = 0.29, standard error = 0.19), while imperfect reconstruction of the transmission networks using sequence data results in bias towards the null away from the true VE_I_ of 0.30 (Figure 1). This imperfect reconstruction is due to the identification of multiple potential infectors for each infectee. For example, another infectee infected by the infector of an index case may share the same number of rare variants as the index case and thus be identified in the top cluster of potential infectors. Of the methods using only sequence data, the shared variant approach using deep sequence data and the hybrid approach return results closest to the true value of VE_I_, while the approaches using consensus sequence data alone return estimates closer to the null. The approaches using clustering result in more accurate estimates of VE_I_ (Figure 1) than the methods weighting all possible infectors, or methods using only the most likely infector(s) (Figure S1).

**Figure 1.**
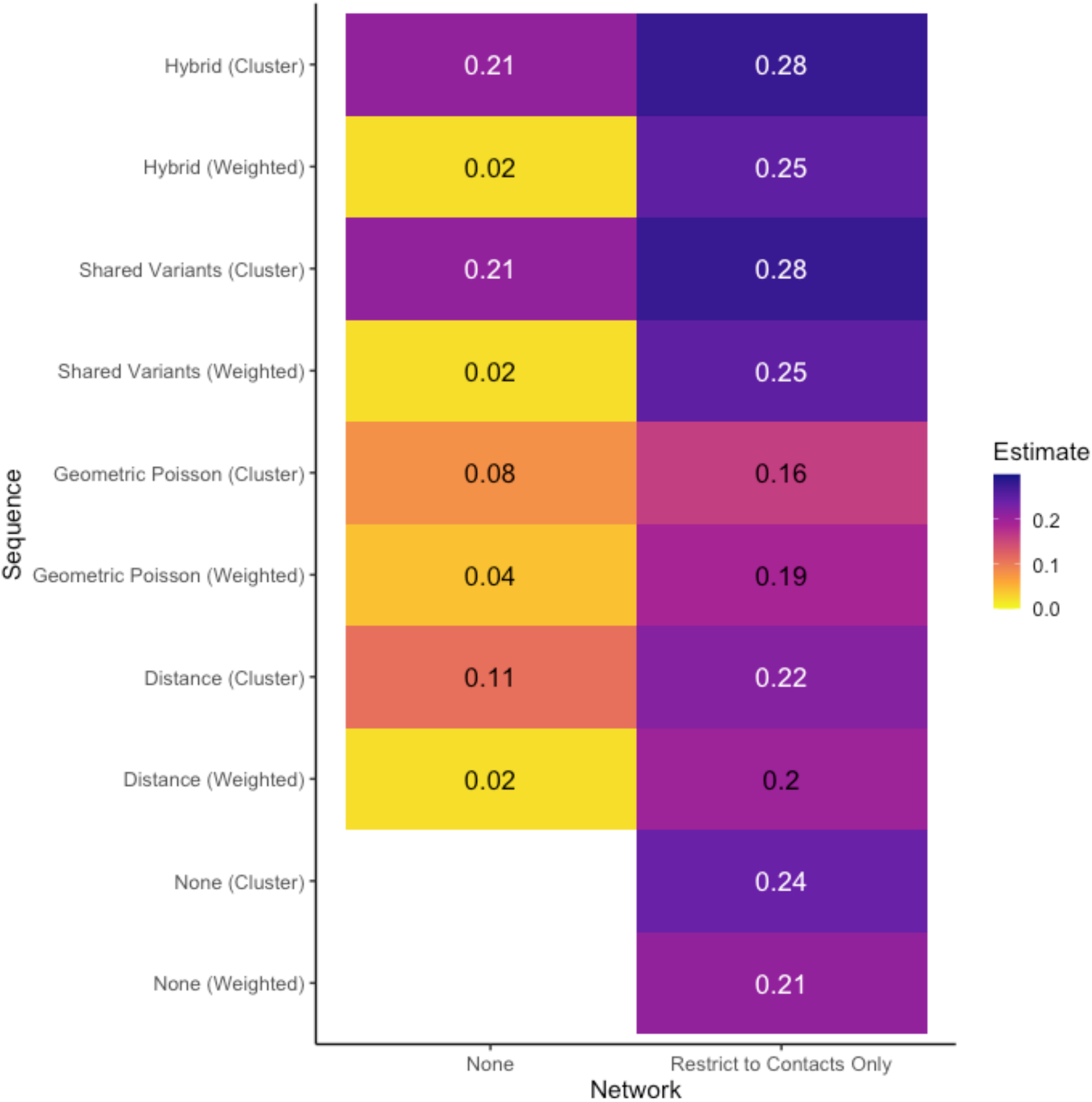
The median VE_I_ estimates from 500 simulations with the baseline parameters, with a true VE_I_ of 0.3. “None” refers to simulations that use only sequence data, without incorporating any epidemiologic information from the network. “Restrict to contacts only” restricts the analysis of sequence data to potential infectors who are contacts from the network.

In reality, sequence data are unlikely to be used in isolation, and adding epidemiologic data from the contact network decreases the bias. Using the infector(s) identified from the hybrid and shared variant approaches among potential infectors restricted to contacts results in median estimates of 0.28, close to the true value of 0.30 (Figures 1 & S1).

The ability to accurately reconstruct transmission networks was previously found to be influenced by parameters such as the bottleneck size and the mutation rate.
^22^
 Varying these and other parameters in our simulations show similar results to the baseline scenario (Figures S2–S5), with the hybrid and shared variant approaches performing worse with a lower mutation rate (Figures S2) and a lower bottleneck size (Figure S3), as expected because less shared variant information is available in these settings.
^22^

## Discussion

In the case of an outbreak of an emerging infectious disease, the ability to rapidly define the efficacy and potential impact of a vaccine is crucial for improving public health and informing policy decisions. An important component of vaccine efficacy which is often overlooked is its ability not only to guard against acquisition of infection by vaccinated individuals, but also to prevent onward transmission from those who are vaccinated that nevertheless become infected. VE_I_ is important for fully understanding and modeling the impact of a vaccine and both sequence and contact tracing data have the potential to allow us to estimate VE_I_ in large individually-randomized controlled trials conducted during an epidemic. Previously, this estimate was only attainable from household and partner studies.^
6–8
^ Advance planning and understanding of the data requirements necessary are critical for obtaining efficacy estimates during the uncertain and urgent setting of an outbreak.

We find that while sequence and contact tracing data have the potential for enabling estimation of VE_I_, misclassification of the trial status of infectors due to imperfect reconstruction of the transmission network leads to bias towards the null of VE_I_ estimates and overall limits our ability to detect an effect of the vaccine on infectiousness. Given the consistent direction of the bias, if an estimate is obtained in a trial using the methods described here, it is expected to be an underestimate of the true VE_I_. The approaches using the top cluster of most likely infector(s) identified from the deep sequence shared variants and hybrid data perform the best of all of the methods using sequence data alone and remain the most accurate method when contact tracing data are incorporated. If deep sequencing data are not available, relying on contact tracing data becomes even more important. The substantial improvement in the estimates when restricting to contacts further underscores the importance of contact tracing for reconstructing transmission networks.

Previous work has pointed to the potential of shared variants identified in deep sequence data, to inform transmission.^
22–24
^ The intuition of this approach is that the pathogen population within an infected host is not composed of identical genomes, but contains some polymorphisms (depending on the population size and the mutation rate). If the transmission bottleneck is sufficiently large, more than one of these genotypes may be transmitted, and the finding that individuals share the resulting polymorphism is then a likely indication of transmission. The methods described here will therefore have variable efficacy for different pathogens. For example, influenza has a high mutation rate,
^9^
 so there is likely sufficient phylogenetic signal and within host variation to support reconstruction of the transmission network and estimation of VE_I_. Initial genomics analyses of SARS-CoV-2 found a low mutation rate;
^30^
 recently, however, there is evidence of minority variants detectable by deep sequencing,
^31^
 suggesting deep sequencing approaches have the potential to be used in ongoing vaccine trials to estimate VE_I_.

Many simplifying assumptions have been made, which could be relaxed in future work. We assume perfect knowledge of infection and recovery times, allowing us to accurately identify the direction of transmission in infector-infectee pairs; in reality, particularly for pathogens with short incubation periods, the direction of transmission may be less clear. We also assume complete and correct sampling of sequence data (which in turn means that everyone in the community is a participant in the trial, as we assume, or at a minimum is followed up in the trial), full knowledge of the contact network, and complete contact tracing. Approaches such as those in the *TransPhylo* R package could be used to assess where cases are likely missing and the overall proportion of the outbreak that has been sampled.
^32^
 A naïve Bayes approach using additional data on individuals in the trial, such as demographic or geographic covariates, has been shown to improve reconstruction of the transmission network when limited sequence and/or contact tracing data are available.
^33^
 Our methods further absorb the limitations of the *seedy* package, which assumes neutral evolution and does not permit superinfection, although this latter limitation is likely more of a concern for endemic rather than epidemic disease models.

Despite these simplifying assumptions, this work highlights the potential for existing data sources to be used in the midst of an outbreak to estimate a key measure of vaccine efficacy. It further identifies the data sources that will lead to the most accurate estimation and can thus be used for better targeting of the limited resources available for data collection in the midst of an epidemic.

## Data Availability

Code is available on github.

https://github.com/rek160/ADAGIO_WPA

## Acknowledgments

This research is funded by the Department of Health and Social Care using UK Aid funding and is managed by the NIHR. RW was supported by funding from R01 AI136947 from the National Institute of Allergy and Infectious Diseases. SVL was supported by NIHGMS R01GM122876 and T32GM074905. The views expressed in this publication are those of the authors and not necessarily those of the Department of Health and SocialCare or the NIH.

We thank Dr. Laura F. White for helpful discussion.

## Supplement text 1

### Aim

In order to estimate VE_I_, we need to estimate the **ratio** of the # people infected by a vaccinated person to the # infected by a control

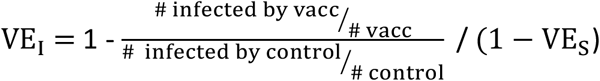

Because # vacc = # cont:

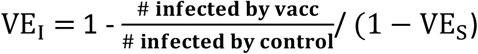

### Analysis

The ratio of the # infected by vacc / # infected by control comes from summing probabilities based on the vaccination status of potential infectors across all infectees.

Example for 1 infected person (probability could be obtained from any of the approaches described in the Methods)

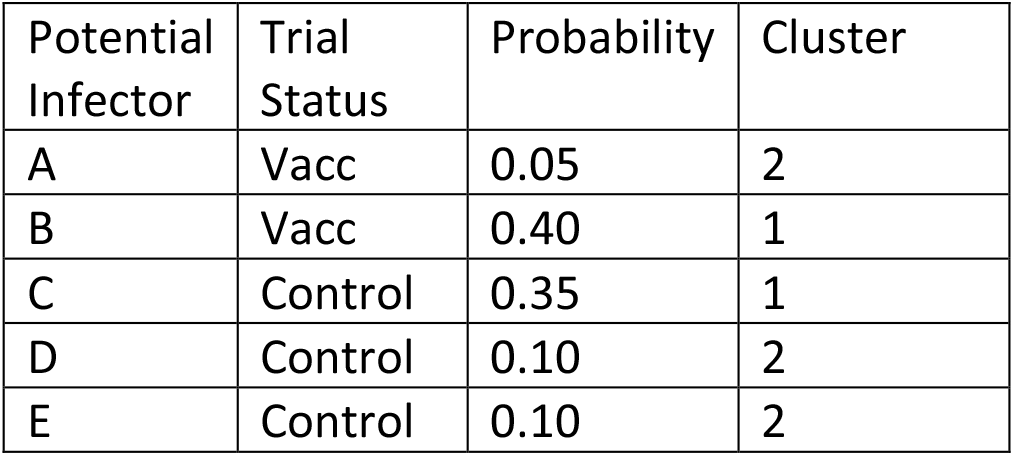

| Potential Infector | Trial Status | Probability | Cluster |
| --- | --- | --- | --- |
| A | Vacc | 0.05 | 2 |
| B | Vacc | 0.40 | 1 |
| C | Control | 0.35 | 1 |
| D | Control | 0.10 | 2 |
| E | Control | 0.10 | 2 |

#### All prob

Include all potential infectors and count each as their probability. We add 0.45 [0.05+0.4] to the numerator of the ratio above and 0.55 [0.35 + 0.10 + 0.10] to the denominator.

#### Cluster

Divide infectors into clusters based on biggest gap in probability (here: 0.35-0.10 = 0.25). Include top cluster if gap is greater than or equal to the threshold (in simulations above, threshold= 0.2 so 0.25 meets this criteria). We add 0.40/(0.40+0.35) to the numerator of the ratio above and 0.35/(0.40+0.35) to the denominator because only 0.40 and 0.35 fall into the top cluster.

#### Max

Include only potential infector(s) that have the highest probability and count as 1. Here, we would add 1 to the numerator of the ratio and nothing to the denominator because 0.4, the maximum probability, is a vaccinated person. If two are tied for most likely then each counts ½.

**Figure S1.**
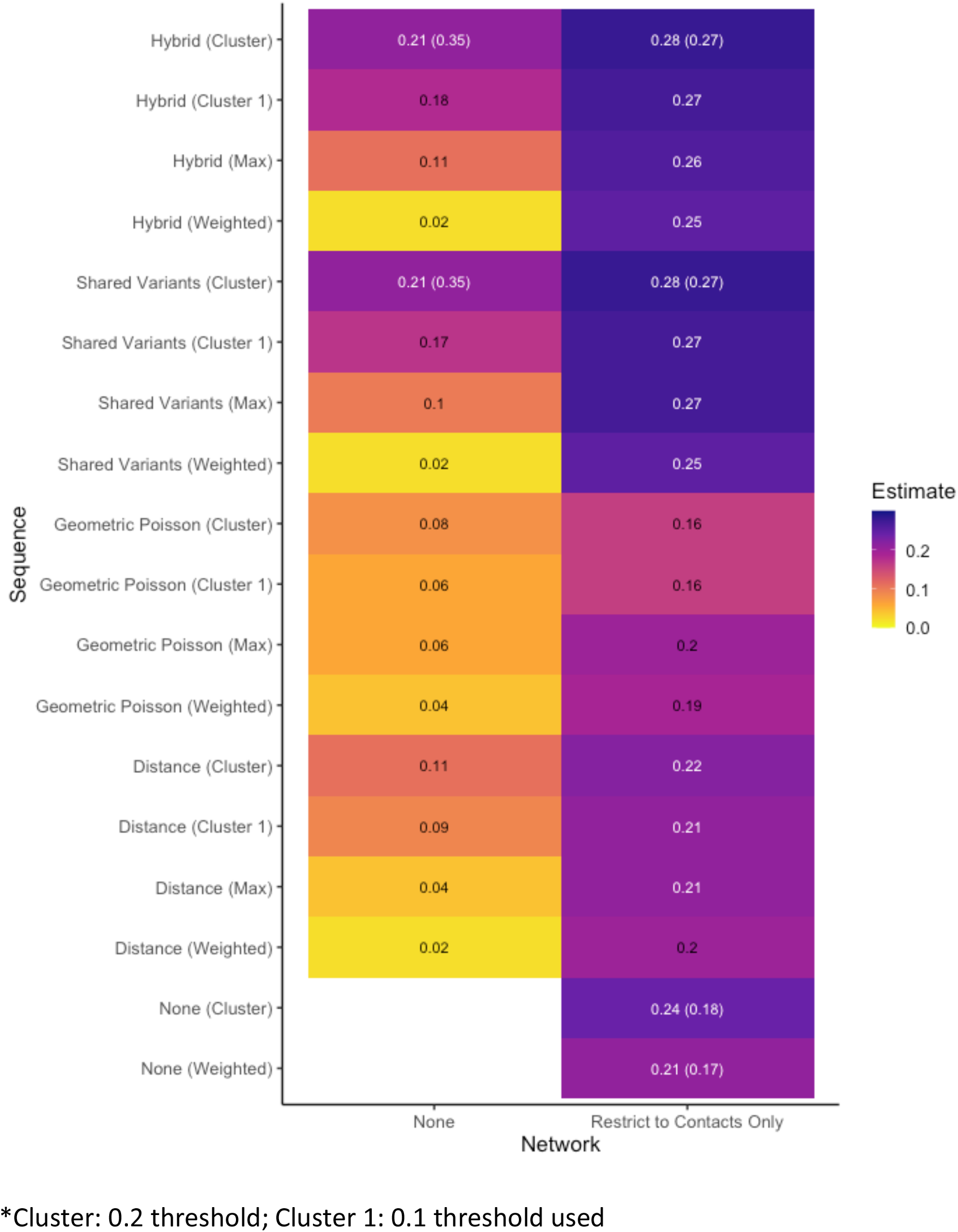
All methods. The median VE_I_ estimates from 500 simulations with the baseline parameters, with a true VE_I_ of 0.3. All approaches, including multiple clustering thresholds and the approach using the vaccination status of only the most likely infector(s) (“Max”), are shown, as well as the average standard errors for the best performing approaches.

**Figure S2.**
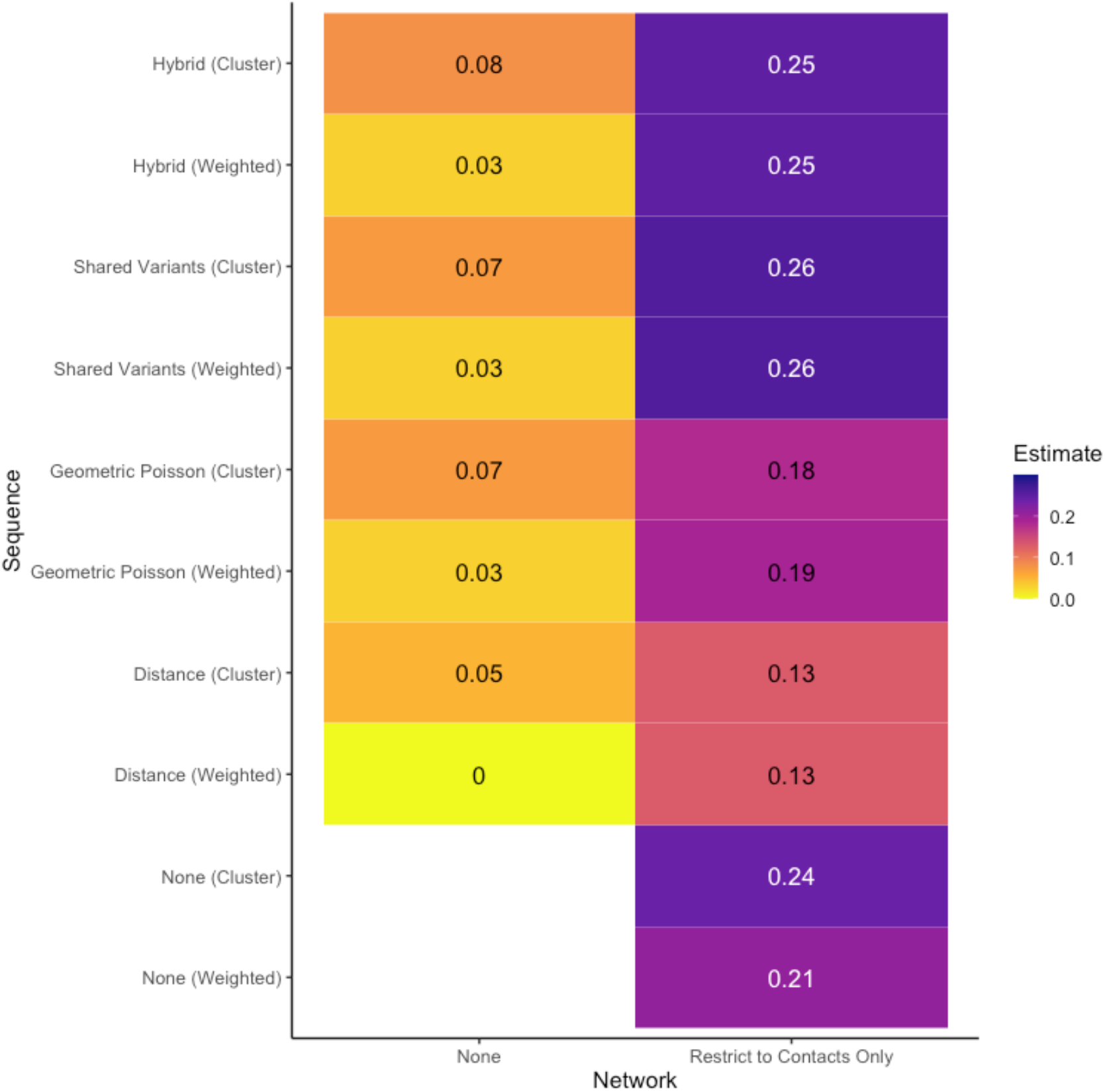
Mutation rate = 0.003. The median VE_I_ estimates from 500 simulations with a lower mutation rate of 0.003, with a true VE_I_ of 0.3.

**Figure S3.**
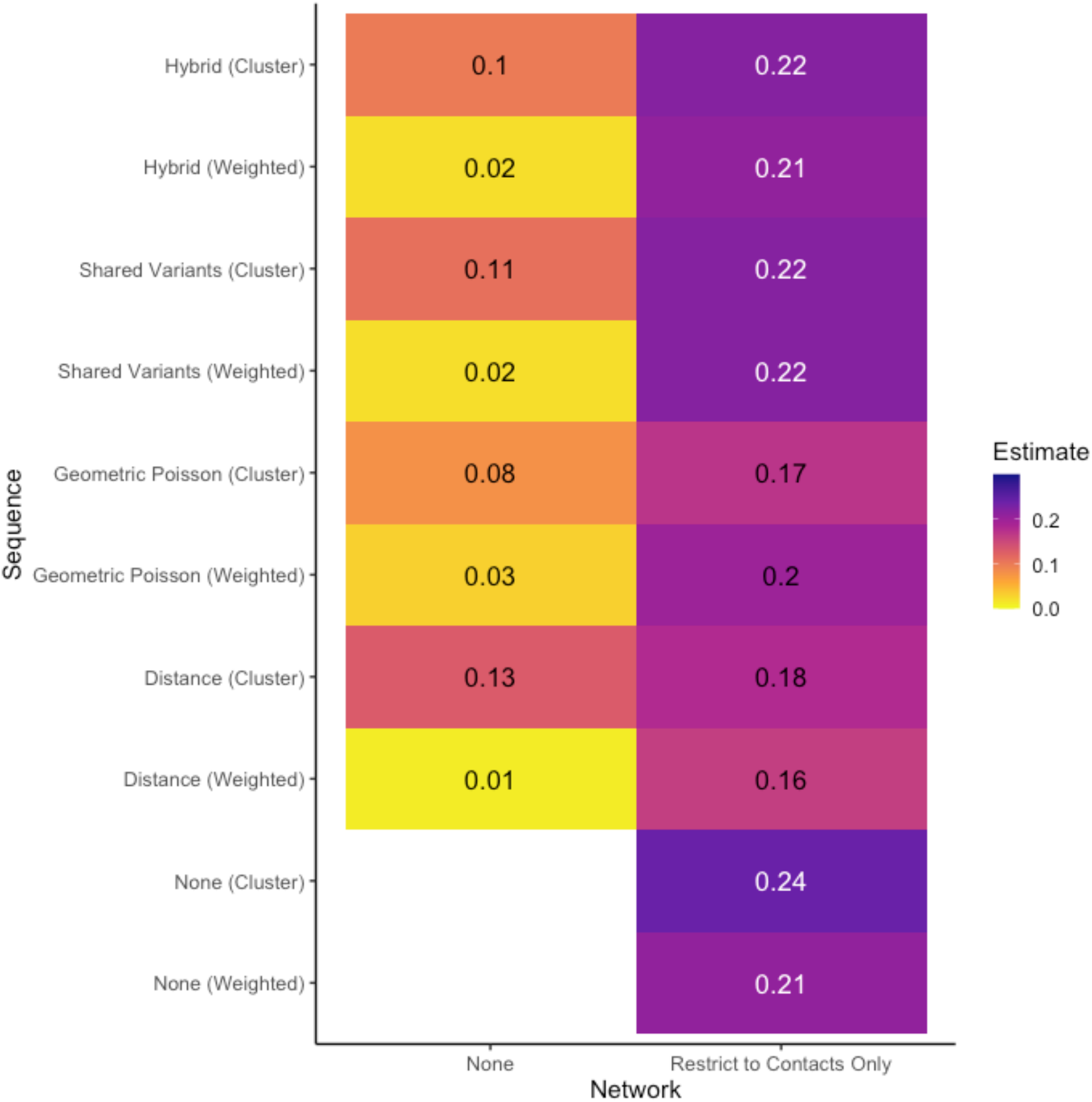
Bottleneck = 2. The median VE_I_ estimates from 500 simulations with a lower bottleneck size of 2, with a true VE_I_ of 0.3.

**Figure S4.**
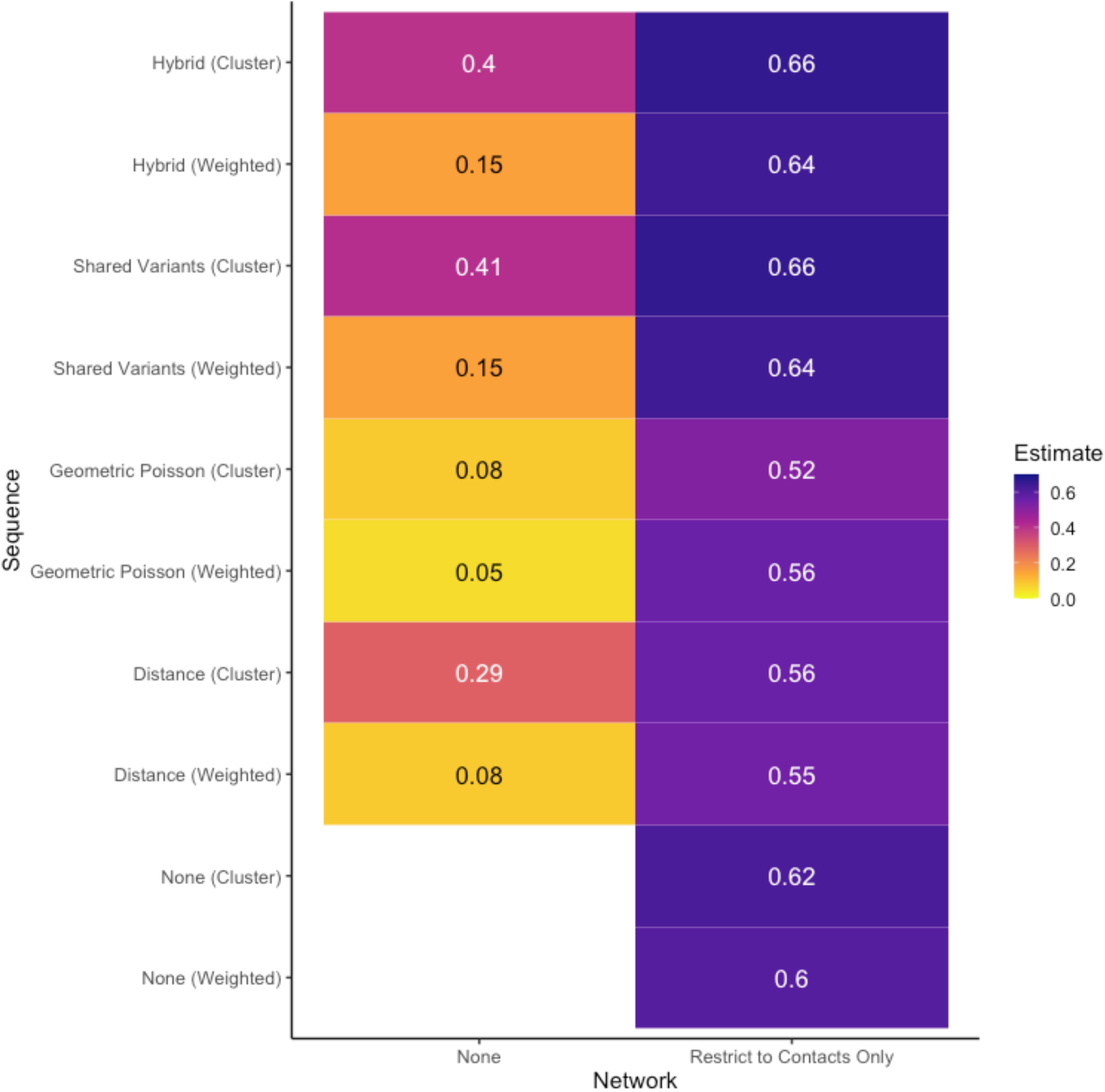
VE_I_ = 0.7. The median VE_I_ estimates from 500 simulations, with a true VE_I_ of 0.7.

**Figure S5.**
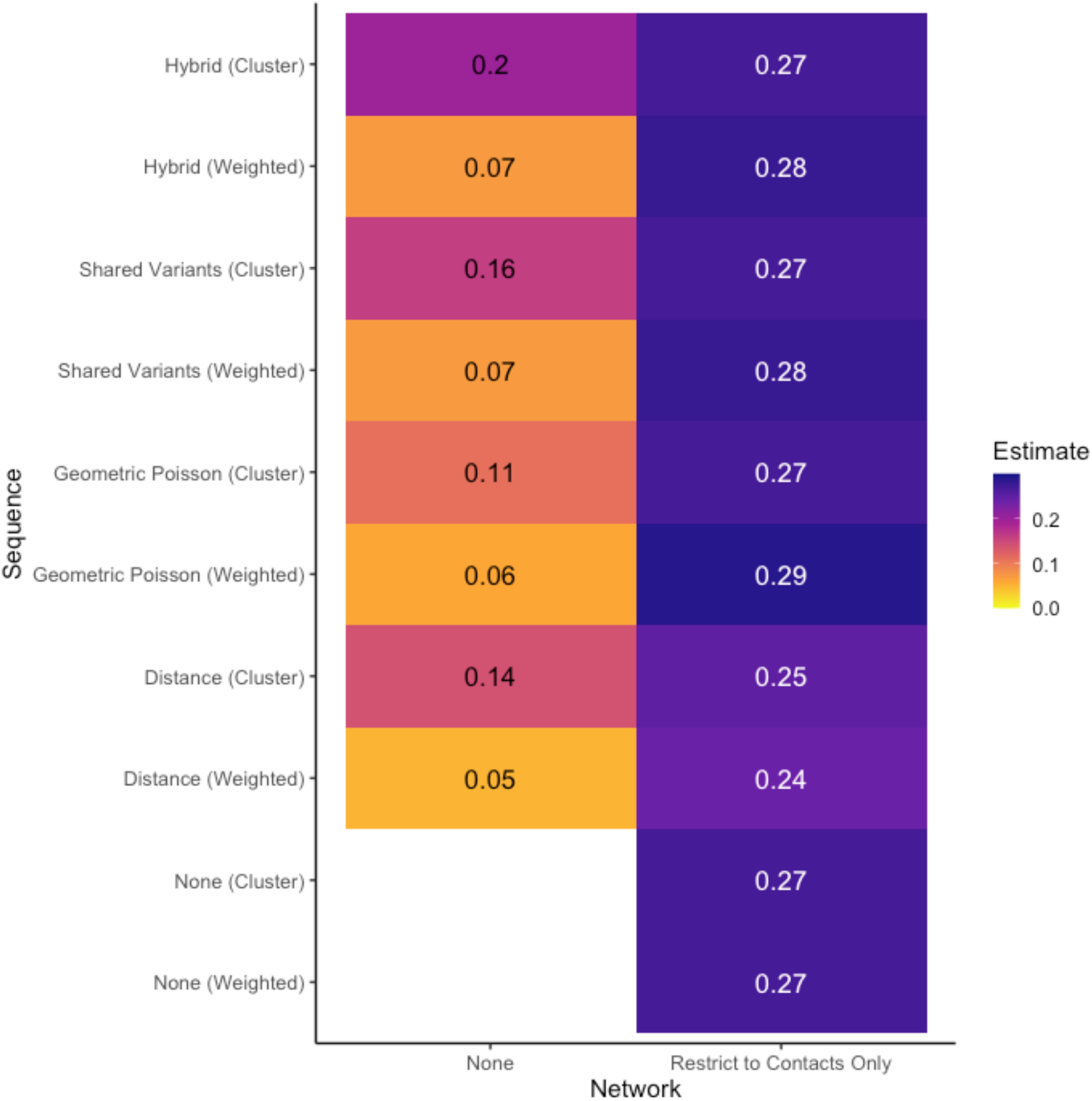
VE_S_ = 0.8. The median VE_I_ estimates from 500 simulations with a VE_S_ of 0.8, with a true VE_I_ of 0.3.

